# Childhood Inflammatory Markers and Risks for Psychosis and Depression at Age 24: examination of temporality and specificity of association in a population-based prospective birth cohort

**DOI:** 10.1101/2020.11.25.20238436

**Authors:** Benjamin I. Perry, Stanley Zammit, Peter B. Jones, Golam M. Khandaker

## Abstract

**Background:** Cross-sectional studies have reported elevated concentrations of inflammatory markers in psychosis and depression. However, questions regarding temporality and specificity of association, crucial for understanding the potential role of inflammation, remain.

**Methods:** Based on 2,224 ALSPAC birth cohort participants, we used regression analyses to test associations of interleukin-6 (IL-6) and C-reactive protein (CRP) levels at age 9 with risks for psychosis (psychotic experiences; negative symptoms; psychotic disorder), and depression (depressive episode; symptom score) at age 24. Regression models were adjusted for sex, ethnicity, social class and body mass index. We tested for linearity (using quadratic terms) and specificity (using bivariate probit regression) of association, and used multiple imputation to explore the impact of missing data.

**Results:** After adjustments, higher IL-6 levels at age 9 were associated with increased risk of psychotic disorder (OR=1.56; 95% C.I., 1.10-2.21 per SD increase in IL-6; OR=1.49; 95% C.I., 1.02-2.18 for the top compared with bottom third of IL-6) and depressive episode (OR=1.14; 95% C.I., 0.99-1.32 per SD increase in IL-6; OR=1.49; 95% C.I., 1.02-2.18 for the top compared with bottom third of IL-6). IL-6 was associated with negative symptoms after adjusting for depression (β=0.09; 95% C.I., 0.01-0.22). There was no evidence for outcome-specific associations of IL-6. Childhood CRP was not associated with adult psychosis or depression.

**Conclusions:** Evidence for similar, longitudinal, dose-response associations suggest that elevated childhood IL-6 could be a shared risk factor for psychosis and depression. The IL-6 pathway may represent a novel target for treatment and prevention of these disorders.

## 1. Introduction

There is mounting evidence for an immune/inflammatory component to psychosis and depression (Brown and Derkits, 2010; Karlsson and Dalman, 2020; Khandaker et al., 2015). Meta-analyses of cross-sectional studies have reported increased levels of circulating inflammatory markers in depression (Goldsmith et al., 2016; Haapakoski et al., 2015; Howren et al., 2009; Köhler et al., 2017; Osimo et al., 2019) and psychosis (Fernandes et al., 2016; Karageorgiou et al., 2018; Miller et al., 2011; Potvin et al., 2008; Upthegrove et al., 2014) compared with controls, indicating a potentially transdiagnostic role of inflammation in major mental disorders. However, cross-sectional studies cannot address whether inflammation is a cause or consequence of illness (i.e., reverse causality). Emerging evidence from longitudinal studies, which are better suited to examine direction of association, suggest associations between elevated levels of circulating inflammatory markers at baseline and increased risks for depression and psychosis at follow-up. A previous study from the Avon Longitudinal Study of Parents and Children (ALSPAC) birth cohort reported that higher levels of interleukin (IL-6), a proinflammatory cytokine, at age 9 were associated with risks for depression and psychosis at age 18 years (Khandaker et al., 2014). Similar findings have been reported from other cohorts and population samples (Gimeno et al., 2009; Kappelmann et al., 2019; Lamers et al., 2019; Metcalf et al., 2017; Wium-Andersen et al., 2013; Zalli et al., 2016). While these longitudinal studies go some way to address the issue of direction of association, there are three key outstanding questions regarding the potential role of childhood inflammatory markers in depression and psychosis.

First, most longitudinal studies have typically focused on a single outcome, so cannot test specificity; i.e., whether the association of childhood inflammation with psychosis and depression at follow-up is stronger for one outcome than other, indicating specificity of effect, or is similar between outcomes. Addressing this issue is important since it may elucidate potential unique or common mechanisms for psychosis and depression. Second, previous studies of childhood inflammatory markers and psychotic outcomes at follow-up have typically focused on positive symptoms. Negative symptoms are key components of the psychosis syndrome and phenomenologically similar to depressive symptoms, but it remains unclear whether childhood inflammation is associated with negative symptoms after controlling for current mood. Third, while previous studies have reported associations of childhood inflammatory markers with depressive and psychotic symptoms in adolescence/early-adulthood (Khandaker et al., 2014; Zalli et al., 2016), it remains unclear whether these associations persist into adulthood. This is important because it could extend the temporal association to early adulthood, a period when most cases of psychiatric disorders are likely to emerge.

We used data from the ALSPAC birth cohort to examine temporality and specificity of association of IL-6 and CRP levels at age 9 and risks for psychosis and depression at age 24, addressing some of the key gaps in the literature. As outcomes, for psychosis we used psychotic experiences (PEs), negative symptoms and psychotic disorder. For depression, we used depressive episode and depression severity score. We examined linearity of association, and whether inflammation was a shared or specific risk factor for psychosis and depression. Finally, we conducted sensitivity analyses to assess the robustness of findings; First, we examined whether associations were stronger in moderate/severe depressive episode compared with mild depressive episode; Second, we repeated our main analyses after imputing missing data.

We predicted that childhood inflammatory markers would be associated with risks for psychotic and depression outcomes at follow-up, suggestive of a potential role of childhood inflammation in pathogenesis of adult depression and psychosis.

## 2. Material and Methods

### 2.1 Description of cohort and sample selection

The ALSPAC birth cohort initially recruited 14,541 pregnant women resident in a geographically defined region in southwest England, with expected dates of delivery between 1.4.1991 and 31.12.1992, resulting in 14,062 live births (Boyd et al., 2013; Fraser et al., 2013; Northstone et al., 2019). An additional 913 participants were recruited subsequently. See www.bris.ac.uk/alspac/researchers/data-access/data-dictionary/ for a fully searchable data dictionary. Study data were collected and managed using REDCap electronic data capture tools hosted at University of Bristol (Harris et al., 2019; Harris et al., 2009). Ethical approval for the study was obtained from the ALSPAC Ethics and Law Committee and Local Research Ethics Committees. All participants provided informed consent.

The risk set for the current study comprised 5,081 participants with data for IL-6 and CRP at age 9. Of these, up to 2,224 participants had data on psychosis outcomes, and 2,219 participants had data on depression outcomes at age 24 (See Supplementary Figure 1 for available samples). We excluded participants with CRP levels >10mg/L to minimize potential confounding by current infection/ chronic inflammatory illness.

**Figure 1:**
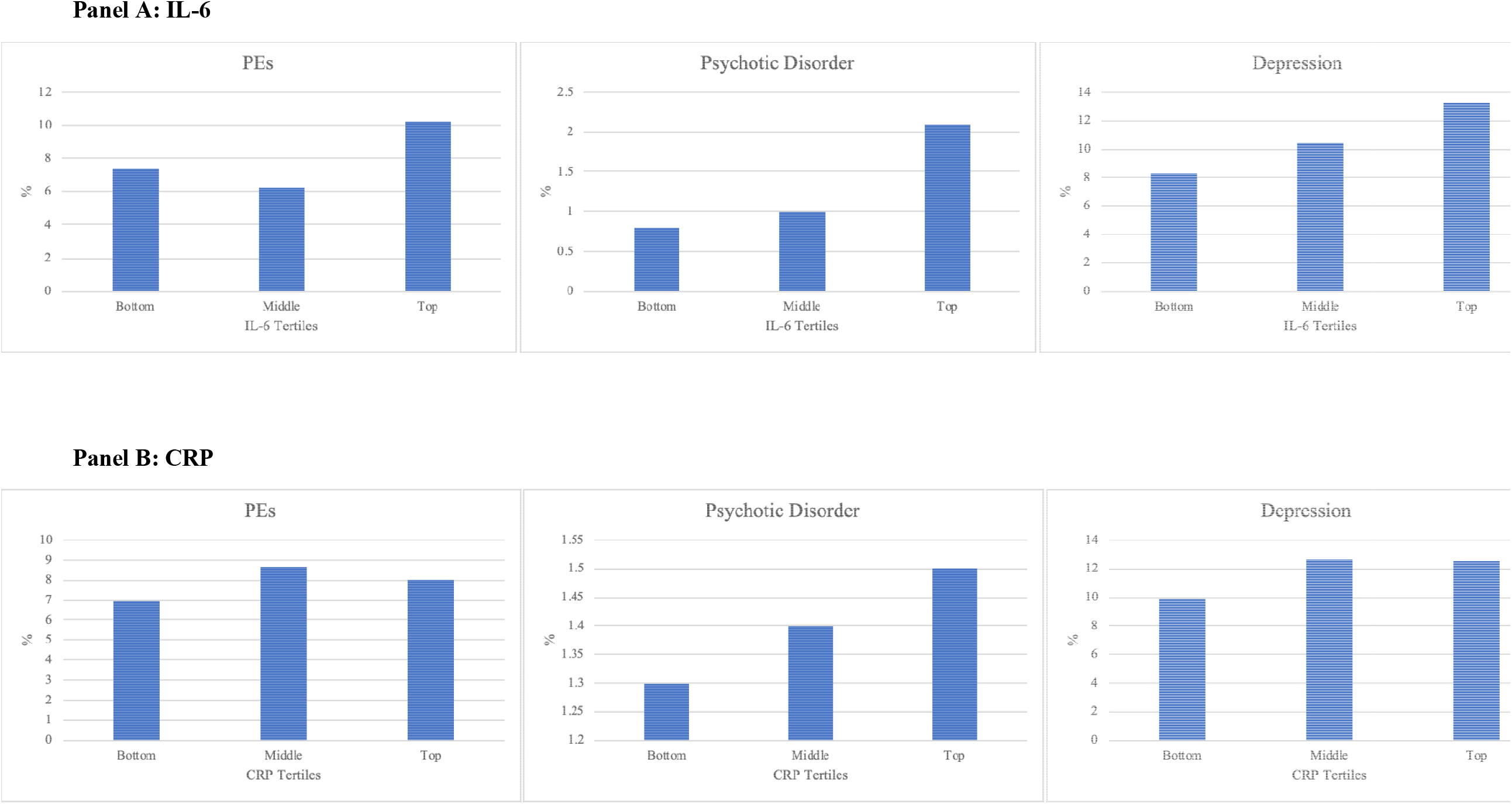
Psychiatric Outcomes at Age 24 per Tertile of IL-6 and CRP Levels at Age 9 in the ALSPAC Birth Cohort.

### 2.2 Psychotic Outcomes at Age 24

#### 2.2.1 Psychotic Experiences (PEs)

PEs were identified through the face-to-face, semi-structured Psychosis-Like Symptom Interview (PLIKSi) conducted by trained psychology graduates and were coded according to the definitions in the Schedules for Clinical Assessment in Neuropsychiatry, Version 2.0. The PLIKSi had good interrater reliability (Intraclass correlation: 0.81; 95% CI, 0.68-0.89) and test-retest reliability (0.9; 95% CI 0.83-0.95) (Sullivan et al., 2020). PEs, occurring in the last six months, covering the three main domains of positive psychotic symptoms were elicited: hallucinations (visual and auditory), delusions (spied on, persecuted, thoughts read, reference, control, grandiosity, and other), and thought interference (insertion, withdrawal, and broadcasting). After cross-questioning, interviewers rated PEs as not present, suspected, or definitely present. Our primary outcome was cases of *definite* PEs; the comparator group was suspected/ no PEs, to maximize the specificity of the outcome variable.

#### 2.2.2 Psychotic Disorder

Cases of psychotic disorder were defined (Sullivan et al., 2020) as having interviewer-rated definite PEs that were not attributable to the effects of sleep/fever, had occurred regularly at least once per month over the previous 6 months, and were either (i) very distressing, (ii) negatively impacted social/occupational functioning, or (iii) led to help-seeking from a professional source. In addition, we included participants meeting the criteria for Comprehensive Assessment of At-Risk Mental State (CAARMS) (Yung et al., 2005) psychotic disorder (threshold psychotic symptoms present for >1 week).

#### 2.2.3 Negative Psychotic Symptoms

Negative symptoms were assessed at age 24 using a questionnaire featuring ten questions based on items from the Community Assessment of Psychic Experiences (CAPE) (Stefanis et al., 2002). Questions covered difficulties with interest, motivation, emotional reactivity, pleasure, and sociability. Participants rated each item on a 4-point scale (0=never; 1=sometimes; 2=often; and 3=always). We were interested in symptoms that were more frequent, thus potentially clinically relevant. Therefore, we recoded each item into a binary variable by coding ‘always’ and ‘often’ as 1=symptom present; ‘never’ and ‘sometimes’ as 0=symptom absent. A total score was constructed by summing ten items (range 0-10).

### 2.3 Depression Outcomes at Age 24 Years

Depression was measured using the computerised version of the Clinical Interview Schedule– Revised (CIS-R), a widely used standardized self-administered tool for measuring depression and anxiety in community samples (Lewis et al., 1992). The CIS-R assesses symptoms of depression occurring in the past week and provides a diagnosis of depressive episode (mild, moderate or severe) based on the *International Statistical Classification of Diseases, 10th Revision* (ICD-10) criteria, which was used as the primary outcome (ICD-10 codes F32.0/F32.1/F32.2). The CIS-R also provides a total depression severity score (0-21) comprising scores for depressed mood, depressive thoughts, fatigue, concentration, and sleep problems, which we also used as an outcome.

### 2.4 Measurement of IL-6 and CRP at Age 9 Years

Blood samples were collected from non-fasting participants during clinic assessment. Samples were immediately spun, frozen and stored at −80°C. There was no evidence of freeze-thaw cycles during storage. In 2008, IL-6 was measured by ELISA (R&D systems, Abingdon, UK), and high-sensitivity CRP (hsCRP) was measured by automated particle-enhanced immunoturbidimetric assay (Roche UK, Welwyn Garden City, UK). All assay coefficients of variation were <5%. Data on IL-6 and CRP were available for 5,071 and 5,081 participants, respectively. In the total sample, CRP values ranged from 0.01 to 45.17mg/L. Thirty-two subjects had CRP levels >10mg/L and were excluded from analysis.

### 2.5 Assessment of Potential Confounders

We adjusted for sex (binary), ethnicity (White vs other), social class (defined from father’s occupation, coded categorically as per the UK Office of National Statistics classification system: I, II, III non-manual, III manual, IV, V), and body mass index (BMI) (weight (kg) / height (m^2^)) measured at age 9.

### 2.6 Statistical Analysis

Pearson’s Correlation was used to test correlations between IL-6 and CRP levels, which were log-transformed and standardised (Z-transformed). We compared the prevalence of binary psychiatric outcomes at age 24 across tertiles of IL-6/CRP at age 9 using likelihood ratio tests.

#### 2.6.1 Association between Childhood Inflammatory Markers and Adult Psychiatric Outcomes

We examined the associations of IL-6/CRP with binary psychiatric outcomes (PEs, psychotic disorder, depressive episode) using logistic regression, and with continuous outcomes (depression severity score, negative symptom score) using linear regression. Continuous outcomes were log-transformed and standardised (Z-transformed). Inflammatory markers were coded as both continuous and categorical variables (tertiles). Using tertiles, odds ratios (ORs) were calculated for participants in the top and middle thirds compared with the bottom third of IL-6/CRP distributions. Linearity of association was tested by inspection of the ORs over the thirds of the inflammatory marker distribution. For logistic regression analyses, ORs and 95% confidence intervals (CIs) represent the increase in risk of outcome per standard deviation (SD) increase in exposure. Linear regression beta coefficients and 95% CIs represent the increase in outcome (in SD) per SD increase in exposure. All regression models were adjusted for sex, ethnicity, social class and BMI. Furthermore, regression models for depression severity score were adjusted for negative symptom score and *vice versa*. Non-linearity was examined by including a quadratic term in regression models (IL-6/ CRP-squared).

#### 2.6.2 Specificity of Associations for IL-6 and CRP with Psychotic and Depressive Outcomes

We used bivariate probit regression to test for commonality or specificity of the associations of IL-6/CRP with psychotic and depressive outcomes. Probit regression jointly modelled the outcomes of definite PEs/psychotic disorder and depressive episode in relation to IL-6/CRP, and then tested for equality of regression parameters expressing the effect of IL-6/CRP on each outcome using likelihood ratio tests. We compared a model that allowed estimates to differ between outcomes with a model where the exposure effect was constrained to be the same for both outcomes. We converted probit estimates into ORs by multiplying probit parameters by 1.6 (Ntzoufras, 2003).

#### 2.6.3 Sensitivity Analyses using Moderate/Severe Depressive Episode as Outcome

We used multinomial logistic regression to calculate ORs and 95% CIs for the association of childhood inflammatory markers (continuous) with mild vs moderate/severe depressive episode, compared to no depressive episode. A likelihood ratio test was used to compare the effect estimates.

#### 2.6.4 Sensitivity Analyses using Multiple Imputation of Missing Data

We used multiple imputation using chained equations (MICE) (Buuren, 2011) to impute missing data for covariates using the *MICE* package in *R* (Buuren, 2011). MICE is a flexible and practical approach to handling missing data, produces asymptotically unbiased estimates and standard errors, and is asymptotically efficient (White et al., 2011). See Supplementary Data for further information. From 100 imputed datasets, we used linear and logistic regression to examine the associations of IL-6 and CRP at age 9 and psychiatric outcomes at age 24, as above. Analyses were performed individually on all imputed datasets, with estimates pooled using Rubin’s rules (Rubin, 1987).

## 3. Results

### 3.1 Baseline Characteristics of Sample

Of the 2,224 participants with data on childhood inflammatory markers and psychotic outcomes at age 24, 163 met criteria for definite PEs (7.3%) and 30 for psychotic disorder (1.3%). Of the 2,219 participants with data on exposures and depression outcomes at age 24, 214 met ICD-10 criteria for depressive episode (9.6%). See Supplementary Table 1 for baseline characteristics of the sample. IL-6 and CRP levels at age 9 were moderately correlated (r=0.45, *p*<0.001).

**Table 1:** Odds Ratios for Psychosis and Depression (Binary Outcomes) at Age 24 per SD Increase in IL-6 and CRP Levels at Age 9.

| Outcome | Risk Factor | OR (95% C.I.) |  | *p*-value for adjusted model |
| --- | --- | --- | --- | --- |
|  |  | Unadjusted | Adjusted for sex, ethnicity, social class, BMI |  |
| Definite PEs | IL-6 | 1.12 (0.95-1.31) | 1.08 (0.91-1.29) | 0.357 |
|  | CRP | 1.08 (0.93-1.27) | 1.04 (0.87-1.26) | 0.600 |
| Psychotic Disorder | IL-6 | 1.42 (1.06-2.01) | 1.56 (1.10-2.21) | 0.014 |
|  | CRP | 0.95 (0.65-1.37) | 1.06 (0.71-1.60) | 0.284 |
| Depressive Episode | IL-6 | 1.17 (1.02-1.34) | 1.14 (0.99-1.32) | 0.076 |
|  | CRP | 1.10 (0.96-1.26) | 1.05 (0.89-1.23) | 0.563 |

### 3.2 Prevalence of Psychosis and Depression at Age 24 per Tertile of IL-6/CRP at Age 9

The prevalence of all psychiatric outcomes was higher for participants in the top third of IL-6 at age 9 years compared with those in the bottom third (*p-*values of χ^2^ tests <0.05). A similar trend was observed for CRP (Figure 1).

### 3.3 Longitudinal Associations of IL-6 and CRP at age 9 with Psychotic Outcomes at Age 24

Increasing levels of IL-6 (continuous variable) were associated with higher risk of psychotic disorder (adjusted OR=1.56; 95% C.I., 1.10-2.21; *p*=0.014) (Table 1), consistent with a linear dose-response effect (*p-*value for quadratic term = 0.638) (Supplementary Table 2). Using IL-6 as a categorical variable, participants in the top, compared with bottom, third of IL-6 at baseline had higher risk of psychotic disorder at follow-up (adjusted OR=2.60; 95% C.I., 1.04-6.53; *p*=0.031) (Table 2).

**Table 2:**
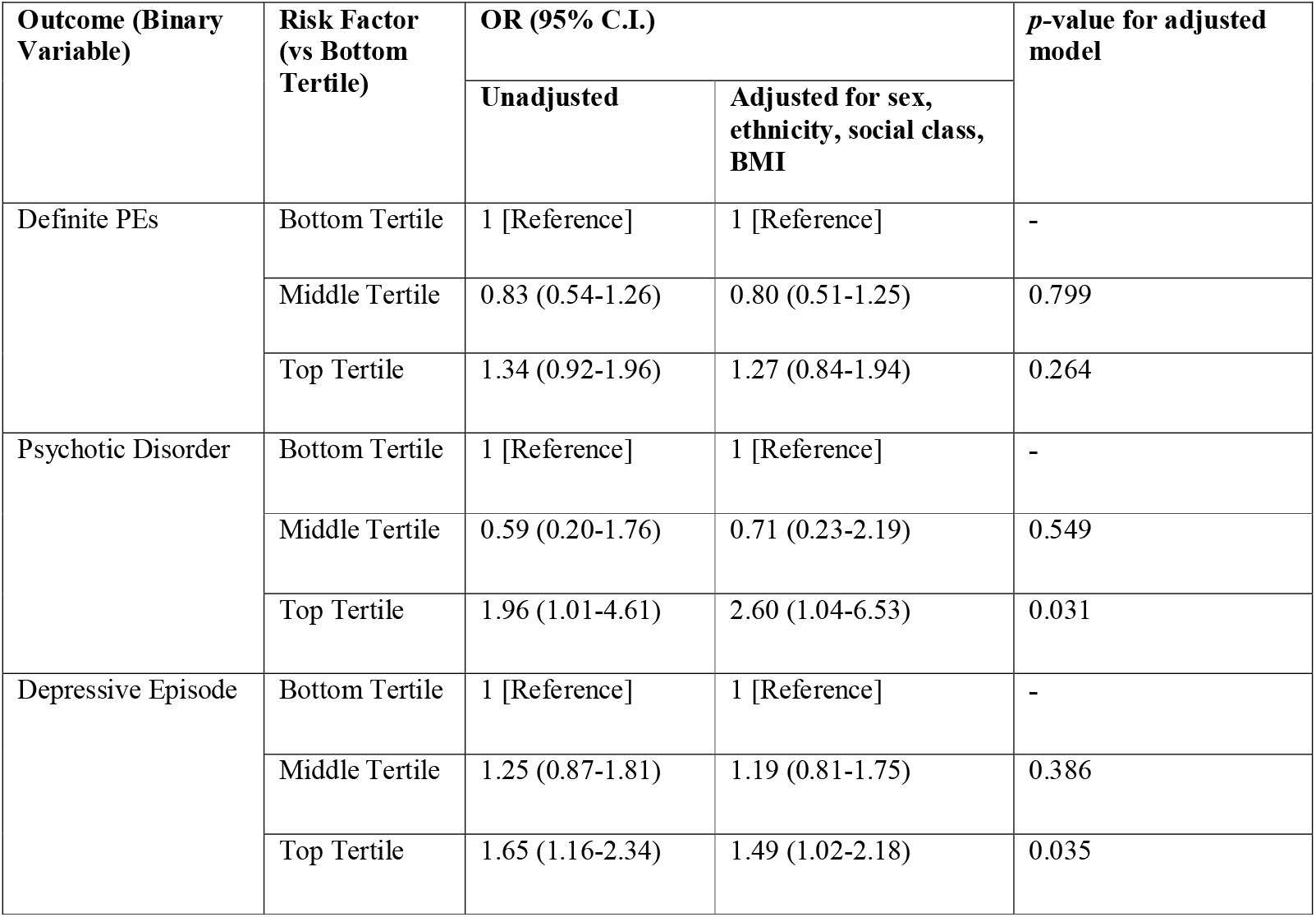
Odds Ratios for Psychosis and Depression (Binary Outcomes) at Age 24 for Participants in Top and Middle Tertiles of IL-6 Distribution Compared with Bottom Tertile at Age 9.

| Outcome (Binary Variable) | Risk Factor (vs Bottom Tertile) | OR (95% C.I.) |  | *p*-value for adjusted model |
| --- | --- | --- | --- | --- |
|  |  | Unadjusted | Adjusted for sex, ethnicity, social class, BMI |  |
| Definite PEs | Bottom Tertile | 1 [Reference] | 1 [Reference] | - |
|  | Middle Tertile | 0.83 (0.54-1.26) | 0.80 (0.51-1.25) | 0.799 |
|  | Top Tertile | 1.34 (0.92-1.96) | 1.27 (0.84-1.94) | 0.264 |
| Psychotic Disorder | Bottom Tertile | 1 [Reference] | 1 [Reference] | - |
|  | Middle Tertile | 0.59 (0.20-1.76) | 0.71 (0.23-2.19) | 0.549 |
|  | Top Tertile | 1.96 (1.01-4.61) | 2.60 (1.04-6.53) | 0.031 |
| Depressive Episode | Bottom Tertile | 1 [Reference] | 1 [Reference] | - |
|  | Middle Tertile | 1.25 (0.87-1.81) | 1.19 (0.81-1.75) | 0.386 |
|  | Top Tertile | 1.65 (1.16-2.34) | 1.49 (1.02-2.18) | 0.035 |

IL-6 levels (continuous variable) at age 9 were associated with increased negative symptoms at age 24 after controlling for potential confounders including current mood (adjusted β=0.09; 95% C.I., 0.01-0.22; *p*=0.041) (Table 3), consistent with a linear effect (*p*-value for quadratic term=0.597) (Supplementary Table 3). Similarly, participants in the top, compared with bottom, third of IL-6 at age 9 had higher negative symptoms (adjusted β=0.20; 95% C.I., 0.04-0.31; *p*=0.021) (Supplementary Table 5).

**Table 3:**
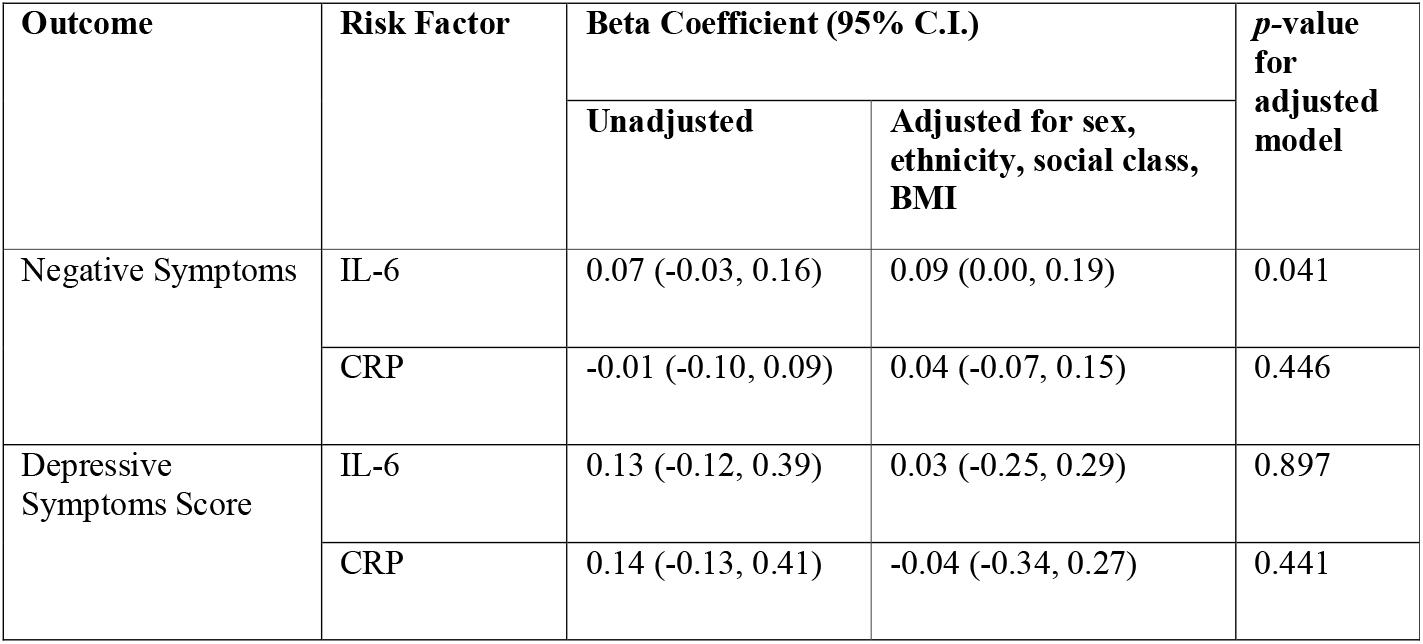
Increase in Negative and Depressive Symptoms (SDs) at age 24 per SD increase in IL-6 and CRP Levels at Age 9.

There was no evidence for an association of IL-6 with PEs (Table 1), or of CRP with any psychotic outcomes (Table 1; Table 3; Supplementary Tables 4 and 6).

**Table 4:** Commonality of Associations for IL-6 and CRP at Age 9 with Psychotic Disorder and Depressive Episode at Age 24.

| Risk Factor | Odds Ratio (95% C.I.) |  |  | LRT<sup>a</sup> Comparing Specific vs. Common Effect |
| --- | --- | --- | --- | --- |
| | Specific Effect on Psychotic Disorder | Specific Effect on Depressive Episode | Common Effect on Both Outcomes | $\chi^2$-statistic, $p$-value<sup>b</sup> |
| IL-6 | 1.29 (1.01-1.65) | 1.13 (1.01-1.28) | 1.16 (1.04-1.30) | $\chi^2=0.93$; 0.336 |
| CRP | 1.01 (0.79-1.30) | 1.07 (0.96-1.21) | 1.07 (0.96-1.19) | $\chi^2=0.28$; 0.597 |

<sup>a</sup>Likelihood Ratio test comparing a model assuming outcome-specific effect for each model vs model where the risk factor is common (i.e. constrained to be the same across outcomes)
<sup>b</sup>Small $p$ -values indicate evidence of differences in fit between the two models, whereby the shared-effect model does not provide adequate fit for the data, and an outcome-specific model provides a better fit.

### 3.4 Longitudinal Associations of IL-6 and CRP at Age 9 with Depression at Age 24

Participants in the top, compared with bottom, third of IL-6 at age 9 had higher risk of depressive episode at age 24 (adjusted OR=1.49; 95% C.I., 1.02-2.18; *p*=0.035) (Table 2). Similarly, participants in the top, compared with bottom third of IL-6 at baseline had higher depression severity score at follow-up (adjusted β=0.43; 95% C.I., 0.01-1.15; *p*=0.049) (Supplementary Table 4). Using IL-6 as a continuous variable, there was weak evidence for an association of IL-6 levels with depressive episode (adjusted OR=1.14; 95% C.I., 0.99-1.32; *p*=0.076) (Table 1), consistent with a linear dose response effect (*p*-value for quadratic term=0.327). However, there was no evidence for an association of IL-6 (continuous variable) with depression severity score at age 24 (Table 3), or of CRP with any depressive outcomes (Table 1; Table 3).

### 3.5 Test for Specificity of Associations of IL-6/CRP with Psychotic and Depressive Outcomes

In bivariate probit regression analysis, we found no evidence that ORs representing outcome-specific associations of IL-6 with psychotic disorder and depressive episode differed from the OR representing a common effect of IL-6 on both outcomes (*p-*value for likelihood ratio test = 0.336), suggesting that IL-6 could be a common risk factor for psychosis and depression (Table 4). Similar results were observed for definite PEs and depressive episode (*p*-value for likelihood ratio test = 0.701) (Supplementary Table 7), and for CRP.

### 3.6 Results for Sensitivity Analyses using ICD-10 Moderate/Severe Depressive Episode as Outcome

Childhood IL-6 levels were more strongly associated with moderate/severe depressive episode (adjusted OR=1.20; 95% C.I., 1.05-1.36), than mild depressive episode (adjusted OR=1.12; 95% C.I., 0.86-1.46) (*p-*value for likelihood ratio test=0.014). There was no evidence for an association of CRP with either mild or moderate/severe depression (Supplementary Table 8).

### 3.7 Results for Sensitivity Analyses using Multiple Imputation for Missing Data

Results were largely similar to primary analyses. Evidence of associations remained for continuous IL-6 levels at age 9 with psychotic disorder (adjusted OR=1.41; 95% C.I., 1.12-2.25) and negative symptoms (adjusted β=0.04; 95% C.I., 0.00-0.07) at age 24. Similarly, following imputation, participants in the top, compared with bottom, third of IL-6 at age 9 had higher risk of psychotic disorder (adjusted OR=1.54; 95% C.I., 1.02-2.11) and higher negative symptoms (adjusted β=0.14; 95% C.I., 0.00-0.29). However, the association of continuous IL-6 levels with depressive episode attenuated slightly (adjusted OR for continuous IL-6 levels=1.08; 95% C.I., 0.98-1.21), (OR for top, compared with bottom third of IL-6=1.19; 95% C.I., 0.98-1.44). As in the primary analysis, there was no evidence for associations of CRP with psychotic or depression outcomes (Supplementary Tables 9-14).

## 4. Discussion

We examined associations of circulating inflammatory markers measured in childhood with risks of psychosis and depression in adulthood in a large population-based prospective birth cohort. We report that raised IL-6 levels in childhood are associated with risks for psychosis and depression at age 24 in a linear dose-dependent fashion. Our results suggest that evidence for associations was stronger in more clinically-relevant outcomes, such as psychotic disorder compared with definite PEs, and moderate/severe depression compared with mild depression. The results remained largely unchanged after imputation of missing data, suggesting that missing data was unlikely to have significantly impacted our findings. To our knowledge, this is one of the first studies to report a longitudinal association between childhood inflammation and negative symptoms in adulthood, independent of concurrent depressive symptoms. Furthermore, having tested outcome-specific vs common effects, we report some of the first evidence for similar associations for IL-6 with psychotic and depressive outcomes, suggesting that childhood IL-6 could be a shared risk factor for adult psychosis and depression.

Our results are consistent with a previous study from the same birth cohort reporting longitudinal, dose-response associations between childhood IL-6 and risks for psychosis and depression at age 18 years (Khandaker et al., 2014). Similar associations have since been reported from other cohorts (Gimeno et al., 2009; Kappelmann et al., 2019; Lamers et al., 2019; Metcalf et al., 2017; Wium-Andersen et al., 2013; Zalli et al., 2016). The current study extends upon previous findings in a number of ways. First, we provide evidence for potential trans-diagnostic role for inflammation, having examined specificity vs. commonality of association between psychosis and depression. Second, we included a larger number of outcomes than in previous studies, particularly negative symptoms which feature less-often in psychosis research. Third, using a longer follow-up, we confirm that the effects of childhood IL-6 on risks for psychosis and depression persist well into adulthood, to a period when most cases of psychiatric disorders emerge.

Our findings support that the association between inflammation and psychiatric disorders transcends traditional diagnostic boundaries (Khandaker et al., 2017). In addition to depression and psychosis, inflammation has been reported to be associated with anxiety (Rossi et al., 2012), post-traumatic stress disorder (Eraly et al., 2014), autism (Brown et al., 2014) and dementias (Schmidt et al., 2002). One explanation for this apparent trans-diagnostic effect could be that inflammation contributes to features common to different syndromes, such as fatigue, anhedonia and cognitive difficulties (Khandaker et al., 2017). However, inflammation is likely to be relevant for some, but not all, cases of depression/ psychosis, because, as with most chronic illnesses, no risk factor alone is necessary or sufficient for causing an illness.

Together with existing evidence, our findings suggest that inflammation could be a common mechanism for a number of commonly comorbid chronic illness, such as depression, schizophrenia, coronary heart disease and diabetes mellitus (Khandaker et al., 2019). These illnesses are associated with inflammation (Danesh et al., 2008; Goldsmith et al., 2016; Pradhan et al., 2001), which could be linked with early-life factors influencing inflammatory regulation, such as impaired foetal development or childhood maltreatment. This idea is consistent with the common-cause or developmental programming hypothesis by David Barker, which posits that exposure to adversity during a critical ‘developmental window’ might permanently alter certain physiologic system(s) leading to increased risk of chronic illnesses in adult life (Barker, 1993). Indeed, childhood adversity, a known risk factor for depression, schizophrenia and cardiovascular disease, is linked with increased levels of circulating inflammatory markers in adulthood (Baumeister et al., 2016). However, since IL-6 was only measured at age 9 in ALSPAC and on one occasion, we are unable to test whether the longitudinal association with adult depression and psychosis is specific to childhood, or with persistently elevated IL-6 levels in childhood, adolescence and adulthood. Future longitudinal studies should seek to include repeat measures of IL-6 over time, to permit a more detailed understanding of its relationship with adult mental disorders.

Although we adjusted for sex, ethnicity, social class and BMI, as with all observational research, residual confounding still might explain the associations of IL-6 with depression and psychosis. Evidence from Mendelian randomization (MR) studies, which use genetic variants regulating levels/activity of a biomarker as proxies to address the issue of confounding, suggest that IL-6 and CRP could be potentially causally related to depression and psychosis (Hartwig et al., 2017; Khandaker et al., 2018b; Khandaker et al., 2019). This would suggest that our results are unlikely to be explained fully by residual confounding. Furthermore, meta-analyses of secondary data from randomised controlled trials (RCTs) suggest that anti-cytokine drugs, including anti-IL-6 drugs, improve depressive symptoms in patients with chronic inflammatory physical illness independently of improving physical illness (Kappelmann et al., 2018; Wittenberg et al., 2019). Our findings, together with existing evidence, support the therapeutic targeting of the immune system in these disorders. However, identifying patients most likely to benefit from immunotherapy would be key for the success of future RCTs, as inflammation is unlikely to be relevant for all patients with depression or psychosis (Miller et al., 2013; Osimo et al., 2019). A recent RCT reported no favourable effect of tocilizumab (anti-IL-6 receptor monoclonal antibody) in patients with established schizophrenia (Girgis et al., 2018), which may be related to patient selection regardless of evidence of immune-activation. In future, further RCTs of immunotherapies including anti-IL-6 treatment for depression and psychosis are required.

One potential explanation for the null findings for CRP could be measurement error, as biomarkers were assayed in non-fasting blood samples. Non-fasting samples may be susceptible to diurnal variation in some cytokines (de Jager et al., 2009), increasing the chance of measurement error that is likely to be random in relationship with the outcome. Random measurement error in relation to the outcome may increase the likelihood of null findings. CRP may be particularly susceptible since it lies downstream of IL-6 on the inflammatory pathway. In addition, although observational studies have consistently reported elevated CRP and IL-6 levels in depression and schizophrenia (Goldsmith et al., 2016; Haapakoski et al., 2015; Miller et al., 2011; Upthegrove et al., 2014), MR analysis has reported a protective effect of CRP for schizophrenia (Hartwig et al., 2017). This is puzzling since IL-6 stimulates the production of CRP. However, it is possible that genetic predisposition for decreased CRP levels/activity increase susceptibility to infection leading to increased schizophrenia risk through immune, neurodevelopmental and/or other mechanisms (Hartwig et al., 2017; Khandaker et al., 2018a). In future, genomic methods which are able to consider the effect of multiple inflammatory markers together, such as multivariable MR, may be helpful to examine the effects of genetically predicted levels of CRP taking into account that for IL-6.

Strengths of this work include the longitudinal design, the use of prospectively collected data from a large general population-based birth cohort, and the inclusion of a larger number of outcomes for psychosis and depression than previous studies. We adjusted for important potential confounders including sex, ethnicity, social class and BMI. We further examined the nature of associations using tests for linearity, for example comparing ORs for binary psychiatric outcomes over thirds of inflammatory marker distribution and including quadratic terms in regression models. The use of bivariate probit analysis permitted testing for specificity of association between childhood inflammatory markers and psychiatric outcomes in adulthood. Limitations of the study include missing data, a common issue for prospective cohort studies. However, we assessed robustness of our findings using sensitivity analyses including multiple imputation for missing data, and the findings were largely unchanged. This suggests that missing data was unlikely to have significantly impacted our findings. Secondly, the dataset we used did not have ICD/DSM diagnosis of schizophrenia available, although our psychotic disorder outcome would likely meet a clinical threshold for the consideration of treatment. Whilst our psychosis outcomes lie along the continuum of psychosis, PEs do not exclusively represent psychosis-risk, and have previously been reported to be associated with other mental disorders including anxiety and depression (Varghese et al., 2011).

The significance of negative symptoms in the general population in the absence of schizophrenia remains unclear (Werbeloff et al., 2015). Also regarding negative symptoms, the outcome measure was derived from a self-report questionnaire, rather than assessment by trained researchers/clinicians. This may have affected the accuracy of our measure for negative symptoms. Additionally, We used childhood inflammatory markers measured at a single time-point, so it is unclear to what extent they reflect persistent inflammation. However, it has been reported that IL-6 levels remain relatively stable over at least a three-year period (Knudsen et al., 2008). Finally, we were only able to include two inflammatory markers in our study, since ALSPAC only collected data on CRP and IL-6. future studies should also include a wider range of inflammatory markers, to foster a broader understanding of network-level changes, possibly involving wider inflammatory pathways.

In conclusion, we report evidence for longitudinal, dose-response associations of childhood IL-6 levels with risks for psychosis and depression in adulthood, with the strongest associations for more clinically relevant outcomes. Furthermore, we provide empirical evidence that IL-6 could be a shared risk factor for these disorders. Our findings suggest that the IL-6 pathway could represent a putative novel target for treatment/ prevention of psychosis and depression.

## Data Availability

Data from ALSPAC can be obtained following application to the ALSPAC executive committee

## Acknowledgements

The authors are extremely grateful to all the families who took part in this study, the midwives for their help in recruiting them, and the whole ALSPAC team, which includes interviewers, computer and laboratory technicians, clerical workers, research scientists, volunteers, managers, receptionists and nurses.

## Financial Support

BIP acknowledges funding support from the NIHR (Doctoral Research Fellowship, DRF-2018-11-ST2-018). The views expressed in this publication are those of the author(s) and not necessarily those of the NHS, the National Institute for Health Research or the Department of Health and Social Care. GMK acknowledges funding support from the Wellcome Trust (Intermediate Clinical Fellowship; grant code: 201486/Z/16/Z), the MQ: Transforming Mental Health (Data Science Award; grant code: MQDS17/40), the Medical Research Council UK (MICA: Mental Health Data Pathfinder; grant code: MC_PC_17213 and Therapeutic Target Validation in Mental Health; grant code: MR/S037675/1), and the BMA Foundation (J Moulton grant 2019). PBJ acknowledges funding from the MRC and MQ (as above), programmatic funding from NIHR (RP-PG-0616-20003) and support from the Applied Research Collaboration East of England. SZ is supported by the NIHR Biomedical Research Centre at University Hospitals Bristol NHS Foundation Trust and the University of Bristol. The UK Medical Research Council and Wellcome Trust (Grant no: 102215/2/13/2) and the University of Bristol provide core support for ALSPAC. A comprehensive list of grants funding is available on the ALSPAC website (http://www.bristol.ac.uk/alspac/external/documents/grant-acknowledgements.pdf/); this research was specifically funded by The Wellcome Trust (Grant no: 08426812/Z/07/Z), Wellcome Trust & MRC (Grant no: 076467/Z/05/Z) and MRC (Grant no: MR/M006727/1).

## Conflicts of Interest

None

## Contributorship Statement

The study was conceived by GMK. Analyses were conceived by BIP, SZ, PBJ and GMK, and conducted by BIP. The manuscript was written by BIP and revised by SZ, PBJ and GMK.

## Figure Captions

**Figure 1: Psychiatric Outcomes at Age 24 per Te**r**tile of IL-6 and CRP Levels at Age 9 in the ALSPAC Birth Cohort**

**Panel A: IL-6**

**Panel B: CRP**

Figures illustrate the prevalence of binary psychiatric outcome per third of inflammatory marker distribution at age 9 years in the ALSPAC cohort

## Notes

### Competing Interest Statement

The authors have declared no competing interest.

### Author Declarations

ALSPAC Ethics and Law Committee and Local Research Ethics Committees

