## Supplementary Data for "Childhood Inflammatory Markers and Risks for Psychosis and Depression at Age 24: examination of temporality and specificity of association in a population-based prospective birth cohort"

**Supplementary Methods**

**Supplementary Figure 1: Flowchart of Available Sample Size for Analyses Presented**

**Age 9**

### *n* with data on exposures

*n* invited = 12556

**CRP**

*n* with data *n*=5081

**IL-6**

*n* with data *n*=5071

**Depression Outcomes**

**Psychosis Outcomes**

### *n* with data on exposure and outcome variables

*n* invited = 9997

*n* attended = 4009

*n* invited = 9997

*n* attended = 3990

**CRP**

*n* with data =2219

**IL-6**

*n* with data =2216

**CRP**

*n* with data =2224

**IL-6**

*n* with data =2219

**Psychosis Outcomes**

**Depression Outcomes**

### *n* with data on exposure and outcome and potential confounders variables

CRP

*n*=2051

CRP

*n*=2054

IL6

*n*=2049

IL6

*n*=2051

**Multiple Imputation for Missing Data**

We used multiple imputation using chained equations (MICE) (15) to replace missing data for covariates on the sample of participants who had outcome data available, using the *MICE* package in *R* (15). Covariates were selected for multiple imputation if they had: 1) <40% missing data from the sample of participants with data on the outcome (26); 2) suitable auxiliary variables available to use as ‘indicators of missingness’, to reduce the impact of bias attributed by the risk of data being ‘missing not at random’ (27). Auxiliary variables were selected based upon their correlation with the covariate and the contribution toward reducing the fraction of missing information (28). Multiple imputation of 100 datasets was performed using the *MICE* package in *R* (15).

After discounting auxiliary variables that displayed evidence of multicollinearity, over 115 auxiliary variables were included (see below). Box-and-Whisker and Density plots were used to check similarities of observed and imputed data. The covariates imputed were: IL-6 (age 9); CRP (age 9); BMI (age 9); ethnicity, and paternal social class. Following imputation, the sample size available for analyses of psychosis outcomes was *n=*3,990, and for depression outcomes *n*=4,009.

| **Age** | **Variable** |
| --- | --- |
| 0 | Gestational Age, Birthweight, Edinburgh Post-Natal Depression Scores, Partners Ethnic Group, Grandmothers Ethnic Group, Family History of Cardiovascular Disease, Perinatal Stressful Life Events Scores, Financial Difficulties Score, Smoking in Pregnancy, Paternal Education, Maternal Education |
| 3 | Vocabulary Score, Plurals Score, Past Tense Score, Word Combination Score, Language Score, Intelligibility Score, Communicative Score |
| 5 | Leptin |
| 7 | IGF-1, Triglycerides, High-Density Lipoprotein, Low-Density Lipoprotein, Body Mass Index, Waist Circumference, Heart Rate, Blood Pressure, Reading Score, Spelling Score, Phoneme Task Score |
| 8 | Weschler Intelligence Scale for Children Scores, Bullying Questionnaire Scores |
| 9 | IGF-1, Fasting Insulin, Triglycerides, High-Density Lipoprotein, Fasting Plasma Glucose, Low-Density Lipoprotein, Body Mass Index, Waist Circumference, Heart Rate, Blood Pressure, Glucose Tolerance, Leptin, Adiponectin, Apolipoprotein AI, Apolipoprotein B, Glycated Haemoglobin, Cortisol, Smoking |
| 10 | Body Mass Index, Waist Circumference, Heart Rate, Blood Pressure, Working Memory Scores |
| 11 | IGF-1 |
| 12 | Alcohol Use, Simple Reaction Time, Digit Vigilance, Choice Reaction Time, Continuity of Attention, Smoking, Substance Use, Sleep Quality |
| 13 | IGF-1, Psychotic Experiences, Alcohol Use, Physical Activity |
| 15 | Adiponectin, Fasting Insulin, High Density Lipoprotein, Fasting Plasma Glucose, Low Density Lipoprotein, Triglycerides, Waist Circumference, Heart Rate, Body Mass Index, Blood Pressure, Physical Activity, Alcohol Use, Cannabis Use, C-Reactive Protein, Smoking, Sleep Quality Scores |
| 18 | Fasting Plasma Glucose, Triglycerides, High-Density Lipoprotein, Triglycerides, Low Density Lipoprotein, Waist Circumference, Fasting Insulin, Body Mass Index, Heart Rate, Blood Pressure, Alcohol Use, Cannabis Use, CRP, Smoking, Sleep Quality, Physical Activity |
| 24 | White Blood Cell Count, Neutrophils, Eosinophils, Basophils, Fasting Plasma Glucose, Triglycerides, High Density Liproprotein, Low Density Lipoprotein, Fasting Insulin, Body Mass Index, Waist Circumference, Blood Pressure, Cannabis Use, Smoking, Heart Rate |

**List of Auxiliary Variables for Multiple Imputation**

**Supplementary Table 1: Baseline Characteristics of Sample**

| **Characteristic** | **IL-6 Tertiles** | | |
| --- | --- | --- | --- |
|  | **Bottom** | **Middle** | **Top** |
| **Male Sex, *n* (column %)** | 995 (58.8) | 837 (51.0) | 728 (42.0) |
| **White British Ethnicity, *n* (column %)** | 1545 (98.4) | 1476 (98.2) | 1534 (97.8) |
| **Social Class, *n* (column %)**  I  II  III - non manual & manual  IV & V | 47 (3.1)  422 (26.9)  733 (46.7)  366 (23.3) | 65 (4.3)  502 (33.4)  679 (45.2)  212 (14.1) | 53 (3.4)  507 (32.3)  739 (47.1)  270 (17.2) |
| **BMI at age 9, mean (SD)** | 16.75 (1.91) | 17.53 (2.56) | 18.49 (3.39) |
| **CRP (mg/L) at age 9, mean (SD)** | 0.26 (0.63) | 0.42 (0.90) | 1.69 (1.21) |

IL-6 = Interleukin-6; CRP=C-Reactive Protein.

**Supplementary Table 2: ORs (95% CI) and *p*-values for Quadratic Terms for IL-6 and CRP (IL-6^2^ and CRP^2^) in Logistic Regression Models Testing Associations of IL-6 and CRP Levels at Age 9 and Risks for Psychosis and Depression at Age 24**

| **Outcome (Binary Variable)** | **Quadratic term for risk factor** | **OR (95% C.I.) adjusted for sex, ethnicity, social class and BMI** | ***p-*value for quadratic term** |
| --- | --- | --- | --- |
| Definite PEs | IL-6-squared | 1.04 (0.97-1.12) | 0.300 |
|  | CRP-squared | 1.03 (0.93-1.14) | 0.596 |
| Psychotic Disorder | IL-6-squared | 0.94 (0.71-1.23) | 0.638 |
|  | CRP-squared | 1.11 (0.91-1.36) | 0.295 |
| Depressive Episode | IL-6-squared | 0.95 (0.86-1.05) | 0.327 |
|  | CRP-squared | 1.06 (0.96-1.16) | 0.251 |

**Supplementary Table 3: Beta-coefficients (95% CIs) and *p*-values for Quadratic Terms for IL-6 and CRP (IL-6^2^ and CRP^2^) in the Linear Regression Models Testing Associations of IL-6 and CRP Levels at Age 9 and Risks for Negative and Depressive Symptoms at Age 24**

| **Outcome (Binary Variable)** | **Risk Factor** | **β (95% C.I.)** | ***p-*value for adjusted model** |
| --- | --- | --- | --- |
|  |  | **Adjusted for sex, ethnicity, social class, BMI** |  |
| Negative Psychotic Symptoms | IL-6-squared | -0.01 (-0.01, 0.01) | 0.597 |
|  | CRP-squared | 0.00 (0.00, 0.00) | 0.825 |
| Depressive Symptom Score | IL-6-squared | 0.01 (-0.02, 0.03) | 0.475 |
|  | CRP-squared | 0.00 (-0.01, 0.01) | 0.276 |

**Supplementary Table 4: ORs (95% CI) and *p­-*values for Risk of Psychosis and Depression at Age 24 for Participants in Middle and Top Thirds of CRP Distributions, Compared with Bottom Third, at Age 9**

| **Outcome (Binary Variable)** | **Risk Factor**  **(vs Bottom Tertile)** | **OR (95% C.I.)** | | ***p-*value for adjusted model** |
| --- | --- | --- | --- | --- |
|  |  | **Unadjusted** | **Adjusted for sex, ethnicity, social class, BMI** |  |
| Definite PEs | Bottom Tertile | 1 [Reference] | 1 [Reference] | **-** |
|  | Middle Tertile | 1.27 (0.85-1.90) | 1.31 (0.85-2.01) | 0.217 |
|  | Top Tertile | 1.18 (0.80-1.76) | 1.13 (0.70-1.83) | 0.612 |
| Psychotic Disorder | Bottom Tertile | 1 [Reference] | 1 [Reference] | **-** |
|  | Middle Tertile | 1.00 (0.65-1.56) | 1.04 (0.40-2.66) | 0.942 |
|  | Top Tertile | 1.02 (0.43-2.43) | 1.43 (0.54-3.82) | 0.471 |
| Depressive Episode | Bottom Tertile | 1 [Reference] | 1 [Reference] | **-** |
|  | Middle Tertile | 1.06 (0.74-1.52) | 0.89 (0.61-1.31) | 0.554 |
|  | Top Tertile | 1.03 (0.43-2.43) | 1.00 (0.67-1.49) | 0.990 |

**Supplementary Table 5: Beta Coefficients (95% CI) and *p*-values for the SD Increase in Negative and Depressive Symptoms for Participants in Middle and Top Thirds of IL-6 Distributions, Compared with Bottom Third, at Age 9**

| **Outcome** | **Risk Factor**  **(vs Bottom Tertile)** | **Beta Coefficient (95% C.I.)** | | ***p-*value for adjusted model** |
| --- | --- | --- | --- | --- |
|  |  | **Unadjusted** | **Adjusted for sex, ethnicity, social class, BMI** |  |
| Negative Symptoms | Bottom Tertile | 1 [Reference] | 1 [Reference] | **-** |
|  | Middle Tertile | 0.04 (-0.03, 0.09) | 0.02 (-0.03, 0.06) | 0.304 |
|  | Top Tertile | 0.24 (0.01, 0.48) | 0.20 (0.04, 0.31) | 0.021 |
| Depressive Symptoms | Bottom Tertile | 1 [Reference] | 1 [Reference] | **-** |
|  | Middle Tertile | 0.10 (-0.09, 0.20) | 0.07 (-0.11, 0.23) | 0.522 |
|  | Top Tertile | 0.65 (0.01, 1.31) | 0.43 (0.00, 1.15) | 0.049 |

**Supplementary Table 6: Beta Coefficients (95% CI) and *p*-values for the SD Increase in Negative and Depressive Symptoms for Participants in Middle and Top Thirds of CRP Distributions, Compared with Bottom Third, at Age 9**

| **Outcome** | **Risk Factor**  **(vs Bottom Tertile)** | **Beta Coefficient (95% C.I.)** | | ***p-*value for adjusted model** |
| --- | --- | --- | --- | --- |
|  |  | **Unadjusted** | **Adjusted for sex, ethnicity, social class, BMI** |  |
| Negative Symptoms | Bottom Tertile | 1 [Reference] | 1 [Reference] | **-** |
|  | Middle Tertile | 0.01 (-0.19, 0.21) | 0.02 (-0.20, 0.18) | 0.708 |
|  | Top Tertile | -0.03 (-0.26, 0.20) | 0.09 (-0.18, 0.36) | 0.521 |
| Depressive Symptoms | Bottom Tertile | 1 [Reference] | 1 [Reference] | **-** |
|  | Middle Tertile | 0.05 (-0.22, 0.29) | -0.01 (-0.25, 0.23) | 0.887 |
|  | Top Tertile | 0.32 (-0.34, 0.98) | 0.02 (-0.76, 0.81) | 0.955 |

**Supplementary Table 7: Commonality of Associations for IL-6 and CRP at Age 9 with PEs or Depressive Episode at Age 24**

| **Risk Factor** | **Odds Ratio (95% C.I.)** | | | **LRT^1^ Comparing Specific vs. Common Effect** |
| --- | --- | --- | --- | --- |
|  | **Specific Effect on**  **Definite PEs** | **Specific Effect on**  **Depressive Episode** | **Common Effect on**  **Both Outcomes** | **χ^2^-statistic, *p-*value^2^** |
| IL-6 | 1.10 (0.97-1.25) | 1.14 (1.01-1.28) | 1.12 (1.02-1.23) | χ^2^=0.15; 0.701 |
| CRP | 1.06 (0.93-1.21) | 1.08 (0.97-1.21) | 1.07 (0.98- 1.19) | χ^2^=0.03; 0.853 |

^1^Likelihood Ratio Test comparing a model assuming outcome-specific effect for each model vs model where the risk factor is common (i.e. constrained to be the same across outcomes)

^2^Small *p*-values indicate evidence of differences in fit between the two models, whereby the shared-effect model does not provide

adequate fit for the data and an outcome-specific model provides a better fit.

**Supplementary Table 8: The ORs (95% CI) and *p-*values for Mild and Moderate/Severe Depressive Episode at age 24 per SD Increase in IL-6 and CRP Levels at Age 9**

| **Risk Factor** | **Adjusted Odds Ratio (95% C.I.)** | | |  |
| --- | --- | --- | --- | --- |
|  | **No Depressive Episode** | **Mild Depressive Episode** | **Moderate/Severe Depressive Episode** | **Likelihood Ratio Test (χ^2^, *p*-value)** |
| IL-6 | 1 [Reference] | 1.12 (0.86-1.46) | 1.20 (1.05-1.36) | 9.43, 0.014 |
| CRP | 1 [Reference] | 1.01 (0.76-1.33) | 1.09 (0.92-1.29) | 0.87, 0.648 |

**Supplementary Table 9: Odds Ratios for Psychosis and Depression (Binary Outcomes) at Age 24 per SD Increase in IL-6 and CRP Levels at Age 9 After Multiple Imputation**

| **Outcome** | **Risk Factor** | **OR (95% C.I.)** | | ***p-*value for adjusted model** |
| --- | --- | --- | --- | --- |
|  |  | **Unadjusted** | **Adjusted for sex, ethnicity, social class, BMI** |  |
| Definite PEs | IL-6 | 1.08 (0.94-1.25) | 1.05 (0.90-1.21) | 0.540 |
|  | CRP | 1.07 (0.93-1.23) | 1.00 (0.85-1.17) | 0.924 |
| Psychotic Disorder | IL-6 | 1.30 (1.01-1.80) | 1.41 (1.12-2.25) | 0.012 |
|  | CRP | 0.96 (0.69-1.34) | 0.97 (0.68-1.39) | 0.858 |
| Depressive Episode | IL-6 | 1.12 (1.00-1.27) | 1.08 (0.98-1.21) | 0.091 |
|  | CRP | 1.09 (0.96-1.24) | 0.98 (0.85-1.14) | 0.826 |

**Supplementary Table 10: Odds Ratios for Psychosis and Depression (Binary Outcomes) at Age 24 for Participants in Top and Middle Tertiles of IL-6 Distribution Compared with Bottom Tertile at Age 9 After Multiple Imputation**

| **Outcome (Binary Variable)** | **Risk Factor**  **(vs Bottom Tertile)** | **OR (95% C.I.)** | | ***p-*value for adjusted model** |
| --- | --- | --- | --- | --- |
|  |  | **Unadjusted** | **Adjusted for sex, ethnicity, social class, BMI** |  |
| Definite PEs | Bottom Tertile | 1 [Reference] | 1 [Reference] | **-** |
|  | Middle Tertile | 0.96 (0.66-1.39) | 0.91 (0.63-1.33) | 0.641 |
|  | Top Tertile | 1.23 (0.88-1.73) | 1.14 (0.81-1.62) | 0.453 |
| Psychotic Disorder | Bottom Tertile | 1 [Reference] | 1 [Reference] | **-** |
|  | Middle Tertile | 0.82 (0.32-2.11) | 0.83 (0.32-2.14) | 0.693 |
|  | Top Tertile | 1.82 (0.98-3.03) | 1.54 (1.02-3.11) | 0.035 |
| Depressive Episode | Bottom Tertile | 1 [Reference] | 1 [Reference] | **-** |
|  | Middle Tertile | 1.19 (0.86-1.64) | 1.11 (0.80-1.53) | 0.535 |
|  | Top Tertile | 1.36 (1.00-1.85) | 1.19 (0.98-1.44) | 0.068 |

**Supplementary Table 11: ORs (95% CI) and *p­-*values for Risk of Psychosis and Depression at Age 24 for Participants in Middle and Top Thirds of CRP Distributions, Compared with Bottom Third, at Age 9 After Multiple Imputation**

| **Outcome (Binary Variable)** | **Risk Factor**  **(vs Bottom Tertile)** | **OR (95% C.I.)** | | ***p-*value for adjusted model** |
| --- | --- | --- | --- | --- |
|  |  | **Unadjusted** | **Adjusted for sex, ethnicity, social class, BMI** |  |
| Definite PEs | Bottom Tertile | 1 [Reference] | 1 [Reference] | **-** |
|  | Middle Tertile | 1.17 (0.83-1.67) | 1.20 (0.79-1.60) | 0.534 |
|  | Top Tertile | 1.15 (0.82-1.64) | 1.01 (0.69-1.47) | 0.975 |
| Psychotic Disorder | Bottom Tertile | 1 [Reference] | 1 [Reference] | **-** |
|  | Middle Tertile | 0.96 (0.42-2.19) | 0.95 (0.42-2.17) | 0.902 |
|  | Top Tertile | 1.00 (0.46-2.14) | 1.01 (0.44-2.28) | 0.988 |
| Depressive Episode | Bottom Tertile | 1 [Reference] | 1 [Reference] | **-** |
|  | Middle Tertile | 1.03 (0.74-1.45) | 0.92 (0.65-1.30) | 0.644 |
|  | Top Tertile | 1.14 (0.83-1.55) | 0.90 (0.64-1.26) | 0.536 |

**Supplementary Table 12: Increase in Negative and Depressive Symptoms (SDs) at age 24 per SD increase in IL-6 and CRP Levels at Age 9 After Multiple Imputation**

| **Outcome** | **Risk Factor** | **Beta Coefficient (95% C.I.)** | | ***p-*value for adjusted model** |
| --- | --- | --- | --- | --- |
|  |  | **Unadjusted** | **Adjusted for sex, ethnicity, social class, BMI** |  |
| Negative Symptoms | IL-6 | 0.04 (-0.06, 0.13) | 0.04 (0.00, 0.07) | 0.039 |
|  | CRP | 0.00 (-0.09, 0.10) | -0.01 (-0.11, 0.10) | 0.944 |
| Depressive Symptoms Score | IL-6 | 0.10 (-0.06, 0.21) | 0.03 (-0.11, 0.16) | 0.726 |
|  | CRP | 0.03 (-0.11, 0.16) | -0.07 (-0.21, 0.08) | 0.359 |

**Supplementary Table 13: Beta Coefficients (95% CI) and *p*-values for the SD Increase in Negative and Depressive Symptoms for Participants in Middle and Top Thirds of IL-6 Distributions, Compared with Bottom Third, at Age 9, After Multiple Imputation**

| **Outcome** | **Risk Factor**  **(vs Bottom Tertile)** | **Beta Coefficient (95% C.I.)** | | ***p-*value for adjusted model** |
| --- | --- | --- | --- | --- |
|  |  | **Unadjusted** | **Adjusted for sex, ethnicity, social class, BMI** |  |
| Negative Symptoms | Bottom Tertile | 1 [Reference] | 1 [Reference] | **-** |
|  | Middle Tertile | -0.01 (-0.23, 0.21) | -0.01 (-0.22, 0.21) | 0.939 |
|  | Top Tertile | 0.13 (-0.08, 0.34) | 0.14 (0.00, 0.29) | 0.023 |
| Depressive Symptoms | Bottom Tertile | 1 [Reference] | 1 [Reference] | **-** |
|  | Middle Tertile | 0.17 (-0.14, 0.49) | 0.11 (-0.20, 0.43) | 0.493 |
|  | Top Tertile | 0.22 (-0.09, 0.53) | 0.12 (-0.20, 0.43) | 0.470 |

**Supplementary Table 14: Beta Coefficients (95% CI) and *p*-values for the SD Increase in Negative and Depressive Symptoms for Participants in Middle and Top Thirds of CRP Distributions, Compared with Bottom Third, at Age 9, After Multiple Imputation**

| **Outcome** | **Risk Factor**  **(vs Bottom Tertile)** | **Beta Coefficient (95% C.I.)** | | ***p-*value for adjusted model** |
| --- | --- | --- | --- | --- |
|  |  | **Unadjusted** | **Adjusted for sex, ethnicity, social class, BMI** |  |
| Negative Symptoms | Bottom Tertile | 1 [Reference] | 1 [Reference] | **-** |
|  | Middle Tertile | 0.09 (-0.23, 0.41) | 0.03 (-0.26, 0.28) | 0.754 |
|  | Top Tertile | 0.13 (-0.18, 0.43) | 0.02 (-0.25, 0.27) | 0.845 |
| Depressive Symptoms | Bottom Tertile | 1 [Reference] | 1 [Reference] | **-** |
|  | Middle Tertile | -0.05 (-0.27, 0.17) | -0.01 (-0.33, 0.31) | 0.955 |
|  | Top Tertile | -0.02 (-0.23, 0.20) | -0.07 (-0.40, 0.27) | 0.697 |
